# Targeting the Coronavirus Nucleocapsid Protein through GSK-3 Inhibition

**DOI:** 10.1101/2021.02.17.21251933

**Authors:** Xiaolei Liu, Anurag Verma, Gustavo Garcia, Holly Ramage, Rebecca L. Myers, Anastasia Lucas, Jacob J. Michaelson, William Coryell, Arvind Kumar, Alexander W. Charney, Marcelo G. Kazanietz, Daniel J Rader, Marylyn D Ritchie, Wade H. Berrettini, David C. Schultz, Sara Cherry, Robert Damoiseaux, Vaithilingaraja Arumugaswami, Peter S. Klein

## Abstract

The coronaviruses responsible for severe acute respiratory syndrome (SARS-CoV), COVID-19 (SARS-CoV-2), Middle East respiratory syndrome (MERS-CoV), and other coronavirus infections express a nucleocapsid protein (N) that is essential for viral replication, transcription, and virion assembly. Phosphorylation of N from SARS-CoV by glycogen synthase kinase 3 (GSK-3) is required for its function and inhibition of GSK-3 with lithium impairs N phosphorylation, viral transcription, and replication. Here we report that the SARS-CoV-2 N protein contains GSK-3 consensus sequences and that this motif is conserved in diverse coronaviruses, raising the possibility that SARS-CoV-2 may be sensitive to GSK-3 inhibitors including lithium. We conducted a retrospective analysis of lithium use in patients from three major health systems who were PCR tested for SARS-CoV-2. We found that patients taking lithium have a significantly reduced risk of COVID-19 (odds ratio = 0.51 [0.35 - 0.74], p = 0.005). We also show that the SARS-CoV-2 N protein is phosphorylated by GSK-3. Knockout of *GSK3A* and *GSK3B* demonstrates that GSK-3 is essential for N phosphorylation. Alternative GSK-3 inhibitors block N phosphorylation and impair replication in SARS-CoV-2 infected lung epithelial cells in a cell-type dependent manner. Targeting GSK-3 may therefore provide a new approach to treat COVID-19 and future coronavirus outbreaks.

**Significance:** COVID-19 is taking a major toll on personal health, healthcare systems, and the global economy. With three betacoronavirus epidemics in less than 20 years, there is an urgent need for therapies to combat new and existing coronavirus outbreaks. Our analysis of clinical data from over 300,000 patients in three major health systems demonstrates a 50% reduced risk of COVID-19 in patients taking lithium, a direct inhibitor of glycogen synthase kinase-3 (GSK-3). We further show that GSK-3 is essential for phosphorylation of the SARS-CoV-2 nucleocapsid protein and that GSK-3 inhibition blocks SARS-CoV-2 infection in human lung epithelial cells. These findings suggest an antiviral strategy for COVID-19 and new coronaviruses that may arise in the future.

## Introduction

COVID-19 is exacting a severe toll on personal and community health, healthcare systems, and the global economy. The response to this crisis will require multiple approaches for detection, prevention, and treatment. With three major betacoronavirus epidemics in less than 20 years, it would also be prudent to anticipate new coronavirus outbreaks in the future. In addition to development of effective vaccines, antiviral strategies that target conserved mechanisms in coronavirus replication and transmission may be needed for COVID-19 and potential future coronavirus outbreaks. Recent high throughput screens have identified bioactive compounds that impair viral replication and infectivity in tissue culture models of infection by the severe acute respiratory syndrome coronavirus-2 (SARS-CoV-2)(1–5). However, their mechanisms of action and their clinical efficacy remain to be fully delineated and additional targets may be needed to combat SARS-CoV-2, new SARS-CoV-2 variants, and potential novel coronavirus outbreaks in the future.

Coronaviruses express a nucleocapsid (N) protein that is essential for viral replication, transcription, and assembly(6–10). N proteins from the JHM strain of mouse hepatitis virus (JHMV) and from SARS-CoV, which caused the 2002-2004 SARS outbreak, are phosphorylated by glycogen synthase kinase-3 (GSK-3) within an arginine-serine (RS) domain present in N proteins of diverse coronaviruses(7-9, 11-14). Phosphorylation of the JHMV N protein is required for recruitment of the RNA helicase DDX1 and for transcription of long subgenomic RNAs(8); inhibition of GSK-3 impairs recruitment of DDX1, binding to viral mRNAs, and viral replication. N protein from SARS-CoV and infectious bronchitis virus (IBV) physically interact with GSK-3(9, 15) and knockdown of *GSK3* impairs IBV replication in Vero cells(15). Furthermore, lithium, which inhibits GSK-3(16) and is widely used in the treatment of bipolar disorder(17, 18), impairs replication of diverse coronaviruses including SARS-CoV, IBV, porcine epidemic diarrhea virus (PEDV), and transmissible gastroenteritis virus (TGEV)(7, 9, 19–21).

These observations suggest that inhibition of GSK-3 could impair coronavirus infections in vivo, including COVID-19(22–24). Recent phosphoproteomic analyses revealed that SARS-CoV-2 N protein is highly phosphorylated within the RS domain(1, 25–27), but whether GSK-3 phosphorylates SARS-CoV-2 N protein and whether lithium has any effect against SARS-CoV-2 have not yet been tested. Here we show that GSK-3 is essential for phosphorylation of the SARS-CoV-2 N protein, that alternative GSK-3 inhibitors impair N phosphorylation and SARS-CoV-2 infection in human lung epithelial cells, and that litihum therapy is associated with significantly reduced risk of COVID-19. Targeting GSK-3 may therefore provide an antiviral therapy for COVID-19 and for coronavirus infections that may arise in the future.

## Results

### Phosphorylation of the SARS-CoV-2 nucleocapsid protein by GSK-3 at two conserved consensus sites

The SARS-CoV N protein shares 20–30% sequence identity with the N proteins of other coronaviruses(6), and despite the limited sequence similarity, they each have an arginine-serine rich (RS) domain that lies between N-terminal and C-terminal conserved domains(6). The RS domains of N from SARS-CoV and JHMV include repeated motifs (SXXXS)(9) that are frequently associated with GSK-3 phosphorylation, in which the C-terminal serine is phosphorylated by a priming kinase(28), which then allows GSK-3 to phosphorylate multiple serines or threonines spaced 4 residues apart in the C to N terminal direction (Fig. 1A). The sequence of the RS domain of SARS-CoV-2 N is 90% similar to N from SARS-CoV, and both proteins contain two sets of three SXXXS motifs each (labeled “a” and “b” in Fig. 1A and 1B). While the N protein sequences within the RS domains of other coronaviruses diverge, they retain SXXXS motifs (Fig. 1B). In addition, the fourth serine (presumed priming site) is always preceded by an arginine in the −3 position (S<u>R</u>XX<u>S</u>). GSK-3-dependent phosphorylation of RS domains has also been reported for multiple splicing factors and other RNA binding proteins(29, 30).

**Fig. 1.**
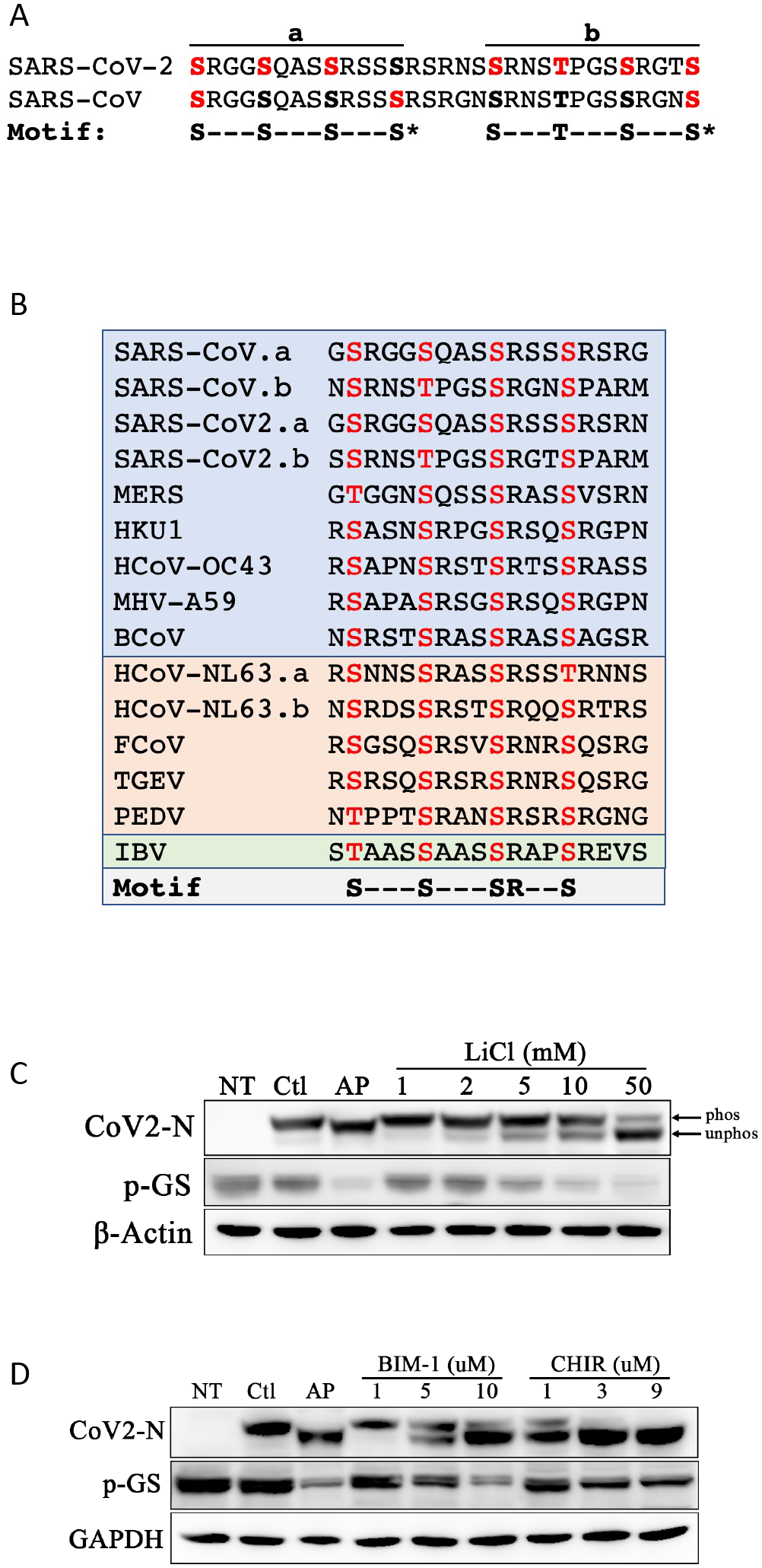
The SARS-CoV-2 Nucleocapsid protein is phosphorylated by GSK-3 in two conserved consensus sites. **A.** The RS domains of SARS-CoV-2 (aa176-206) and SARS-CoV N proteins are 90% identical and contain tandem sets of SXXXS motifs, labelled “a” and “b”. Consensus site serines and threonines are in bold; red indicates sites shown previously by mass spectroscopy to be phosphorylated(1, 9). **B.** Alignment of RS domains in N proteins from pathogenic CoVs showing conservation of repeated SXXXS motifs (“S” in motif represents serine or threonine) and a highly conserved arginine 3 residues before the putative priming sites (S**<u>R</u>**XXS). Blue indicates β-CoV; orange: α-CoV; green: *γ*-CoV. **C.** SARS-CoV-2 N was expressed in 293T cells, treated with LiCl for 18h, and then subjected to SDS-PAGE and immunoblotting for N protein (CoV2-N), phosphorylated glycogen synthase (pGS), or β-Actin as a loading control. Alkaline phosphatase (AP) treatment of cell lysates increases electrophoretic mobility. LiCl inhibits N phosphorylation with IC_50_ ∼10 mM. “phos” indicates phosphorylated N protein; “unphos” indicates dephosphorylated N; NT, nontransfected. Ctl, nontreated control. **D.** SARS-CoV-2 N expressing 293T cells were treated for 18h with bisindolylmaleimide I (BIM-I) or CHIR99021 (CHIR) at the indicated concentrations and cell lysates were immunoblotted as in panel C.

Recent phosphoproteomic analyses have shown that the RS domain of the SARS-CoV-2 N protein is highly phosphorylated(1, 25–27), but whether GSK-3 phosphorylates N protein from SARS-CoV-2 has not been addressed in a cellular context. We expressed SARS-CoV-2 N in human embryonic kidney 293T cells (Fig. 1C) or mouse lung epithelial MLE12 cells (Figure S1A). N phosphorylation was demonstrated by treating cell lysates with alkaline phosphatase, which increased the electrophoretic mobility of N, as observed previously for SARS-CoV N protein(8, 9). Lithium chloride (LiCl) inhibited N phosphorylation with IC_50_ ∼10mM in 293T cells (Fig. 1C, S1B). Phosphorylation of the GSK-3 substrates Glycogen Synthase (GS; Fig. 1C) and ß-catenin (Fig. 2A, lanes 1-3) was also inhibited with an IC_50_ ∼ 10 mM. In contrast, the K_i_ for LiCl inhibition of GSK-3 *in vitro* is 1 mM(31, 32) and the effective in vivo concentration for Li^+^ inhibition of GSK-3 in mice and humans is also 1 mM(33, 34). The relatively high Li^+^ concentration needed to inhibit N phosphorylation ex vivo raises the concern that Li^+^ may act through a target other than GSK-3. To examine this possibility rigorously, we tested multiple, selective GSK-3 inhibitors, including bisindolylmaleimide I (BIM-I), CHIR99021, AR-A014418, and Kenpaullone, all of which inhibited N phosphorylation in the low µM range (Fig. 1D, S1C), strongly supporting that GSK-3 is essential for N protein phosphorylation.

**Fig. 2.**
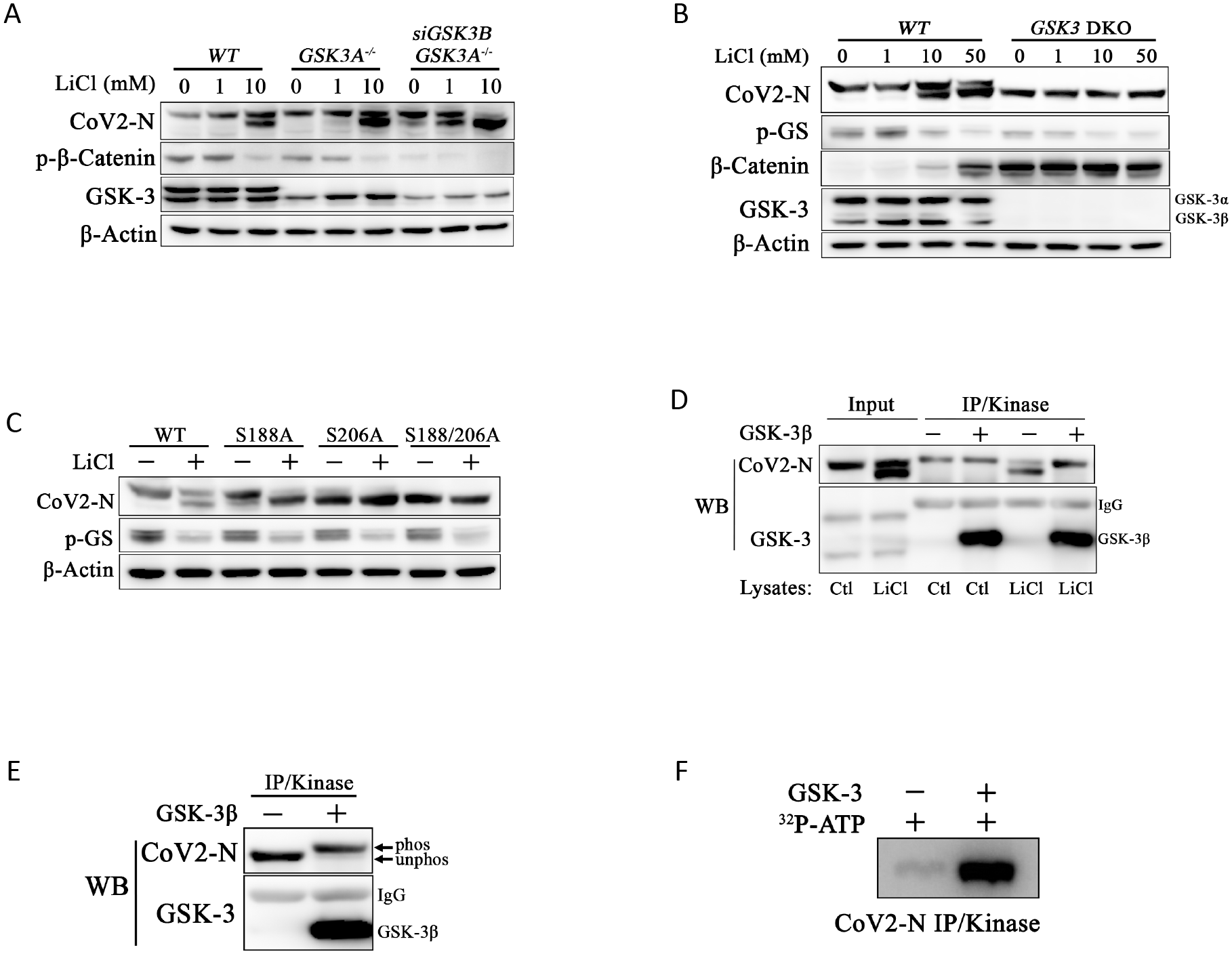
*GSK3* is required for N phosphorylation. **A.** Control 293T cells (*WT*), 293T cells with CRISPR/Cas9 KO of *GSK3A* (*GSK3A^-/-^*), and *GSK3A^-/-^* cells with siRNA knockdown of *GSK3B* (*GSK3A^-/--^*;siGSK3*B*) were treated with LiCl at indicated concentrations and lysates were immunoblotted for N protein, phospho-β-catenin, GSK-3α/β, or β-Actin. Combined loss of *GSKA* and *GSK3B* impairs phosphorylation of N and β-catenin and enhances sensitivity to LiCl. **B.** *GSK3B* was deleted in *GSK3A^-/-^* cells using CRISPR (*GSK3* DKO). N protein was expressed in both wild-type and DKO cells in the presence of increasing concentrations LiC for 18h as above and immunoblotted for N protein, phospho-GS (pGS), total β-catenin, GSK-3α/β, and β-actin. N is not phosphorylated in DKO cells and mobility is not affected by LiCl treatment. Total *β−*catenin protein accumulates in absence of GSK-3 (DKO)(35) or upon inhibition with LiCl(36). **C.** Serine-188 and serine-206 were mutated to alanine by site directed mutagenesis and single and double mutant N proteins were expressed in 293T cells in the presence of vehicle or 10mM LiCl and immunoblotted for N protein, pGS, or β-actin. The double mutant N^S188A;S206A^ migrates similar to dephosphorylated wild-type N. Single mutants are more sensitive to LiCl. **D/E.** N protein was immunoprecipitated from wild-type HEK293T cells treated with or without 10mM LiCl for 18h (indicated by “Ctl” or “LiCl” below each lane in panel D) or from *GSK3* DKO cells (panel E). Immunoprecipitated N protein was added to an *in vitro* kinase reaction with recombinant GSK-3β.GSK-3β phosphorylates N from LiCl treated wild-type and DKO cells as indicated by slower electroporetic mobility (“phos” in panel E). **F.** N protein immunoprecipitated from DKO cells was added to an *in vitro* kinase reaction with recombinant GSK-3β as in panel E except that *γ*-^32^P-ATP was included and gels were fixed, dried, and exposed to X-ray film.

### GSK-3 is required for N phosphorylation

However, these compounds, which inhibit GSK-3 by competing for ATP binding, may have off-target effects. As an alternative and more definitive approach, we used siRNAs and CRISPR/Cas9 to knockdown or knockout (KO) both *GSK3A and GSK3B,* which encode two highly similar GSK-3 isoforms (GSK-3α and GSK-3β, respectively). KO of *GSK3A* alone had a minimal effect on phosphorylation of N or β-catenin (Fig. 2A), consistent with redundant functions of *GSK3A* and *GSK3B*(35). However, siRNA knockdown of *GSK3B* in *GSK3A* KO cells impaired N phosphorylation and increased the sensitivity to LiCl (Fig. 2A), with modest inhibition of N phosphorylation detectable at 1 mM LiCl and clearly enhanced inhibition at 10 mM. Importantly, combined KO of *GSK3A* and *GSK3B* (DKO) completely prevented N protein phosphorylation (Fig. 2B). *GSK3A/B* DKO also reduced GS phosphorylation compared to parental cells and resulted in accumulation of β-catenin protein, which is destabilized by GSK-3 and therefore accumulates when GSK-3 is inhibited or knocked out(35, 36). In the absence of GSK-3, LiCl had no further effect on GS or β-catenin and no effect on N protein mobility, showing that Li^+^ inhibits N phosphorylation solely through inhibition of GSK-3. These pharmacological and genetic data demonstrate unequivocally that GSK-3 is essential for phosphorylation of the SARS-CoV-2 N protein.

GSK-3 substrates that follow the SXXXS motif require a priming phosphorylation at the C-terminal serine or threonine(37, 38); mutation of this residue in established GSK-3 substrates including GS and β-catenin prevents phosphorylation of more N-terminal serines and threonines by GSK-3(37, 39). Similarly, mutation of the two putative priming sites in SARS-CoV N blocks phosphorylation by GSK-3(9). To test whether the SARS-CoV-2 N protein also requires priming site serines, we mutated serine-188 and serine-206 of SARS-CoV-2 N protein to alanines (N^S188A,S206A^) and then expressed the single and double mutant N proteins in HEK293T cells. The single mutant form N^S188A^ migrates in the same position as phosphorylated wild-type N protein whereas the N^S206A^ migrates more rapidly, suggesting that it is hypophosphorylated (Fig. 2C). Mobility of the N^S188A,S206A^ protein is similar to dephosphorylated N protein and is not affected by GSK-3 inhibition LiCl (Fig. 2C) or treatment with alkaline phosphatase (Figure S2A). Additionally, both of the single serine to alanine mutants are more sensitive to LiCl. These data indicate that GSK-3 phosphorylation of SARS-CoV-2 N protein requires the canonical GSK-3 priming site serines.

Although our data show that GSK-3 is required for phosphorylation of N protein at a classical GSK-3 consensus site, it remains formally possible that GSK-3 indirectly regulates N protein phosphorylation. Pharmacological inhibition of GSK-3 also activates mTOR and downstream pathways, including ribosomal protein S6 kinase, elF4E, and SR protein kinase-2 (SRPK2) (40, 41). SRPK1/2 regulate activity of serine/arginine-rich (SR) proteins via phosphorylation of the arginine/serine (RS)-repeat domains. Similar to GSK-3, SRPK1/2 tend to phosphorylate multiple serines within a local domain in a processive manner. SRPK1 can phosphorylate N *in vitro* and has been proposed as a priming protein kinase for GSK-3 phosphorylation of N (4, 12, 26, 42). To determine whether mTOR and SPRK1/2 are involved in N phosphorylation, HEK293T cells expressing SARS-CoV-2 N were treated with the mTOR inhibitor Rapamycin or the SRPK1/2 inhibitor SRPIN340. Rapamycin treatment had no effect on N phosphorylation despite potent inhibition of ribosomal protein S6 kinase phosphorylation (Figure S2B). The SRPK1/2 inhibitor SRPIN340, either alone or combined with LiCl, also did not affect N protein phoshorylation at sites that affect electrophoretic mobility (Figure S2C).

To test whether GSK-3 directly phosphorylates N protein, we performed *in vitro* kinase assays with recombinant GSK-3β and N protein purified by immunoprecipitation from HEK293T cells. Use of N protein expressed in mammalian cells was important because the priming site will not be phosphorylated in bacterially expressed recombinant protein and, without the priming phosphorylation, the GSK-3 sites may not be phosphorylated. HEK293T cells expressing myc-tagged N were treated with the GSK-3 inhibitor LiCl and then N protein was immunoprecipitated. As shown in Fig. 2D, N protein lacking phosphorlyation at the GSK-3 dependent sites migrates more rapidly than phosphorlyated N (lane 5). Recombinant GSK-3β was then added to the immunoprecipitate and the reaction was incubated for 30 minutes. N protein phosphorylated by GSK-3 migrated with slower mobility (Fig. 2D, lane 6), similar to N protein from untreated cells. N protein expressed in *GSK-3* double knock out (DKO) 293T cells, immunoprecipitated, and added to an *in vitro* kinase reaction was completely phosphorylated by recombinant GSK-3β (Fig. 2E). Alternatively, incubation with GSK-3β and *γ*-^32^P-ATP led to robust incorporation of ^32^P into N protein (Fig. 2F), consistent with recent findings from others showing in vitro phosphorylation of the RS domain of N by GSK-3(26, 43). These data demonstrate that GSK-3βdirectly phosphorylates N protein.

### Inhibition of N phosphorylation by clinically well-tolerated GSK-3 inhibitors

The phosphorylation of a motif within an arginine-rich domain, and especially the high conservation of arginine at the −3 position relative to the priming site (Fig. 1B), suggested that the priming kinase may be an arginine-directed, or “basophilic”, protein kinase. We began to test candidate kinases using inhibitors of MAP/ERK kinases (MEK1/2), casein kinase II (CKII), calmodulin-dependent protein kinase II, and protein kinase C (PKC). Although most inhibitors we tested had no effect on N phosphorylation (data not shown), including the PKC inhibitor Gö6976 (Figure S3B), the structurally related PKC inhibitors Enzastaurin, Sotrastaurin, and Gö6983 did inhibit N phosphorylation (Fig. 3A and S3C). Unexpectedly, in addition to potently inhibiting phosphorylation of endogenous PKC substrates induced by the PKC activator PMA (Figure S3A), these bisindolylmaleimides also inhibited phosphorylation of the endogenous GSK-3 substrate GS. To distinguish whether the inhibition of N phosphorylation was due to inhibition of GSK-3 or inhibition of a priming phosphorylation by PKC, we knocked down expression of PKC-α, PKC-*δ*, and PKC-*ε*, the major PKC isoforms expressed in HEK293T cells. Single or combined knockdown of PKC had no effect on phosphorylation of N or GS (Figure S3D). To confirm that Enzastaurin directly inhibits GSK-3, we performed *in vitro* kinase assays. Phosphorylation of the GSK-3 substrate tau and N protein was inhibited by Enzastaurin *in vitro* (Fig. 3B and 3C), confiming that Enzastaurin directly inhibits GSK-3, consistent with a prior report(44).

**Fig. 3.**
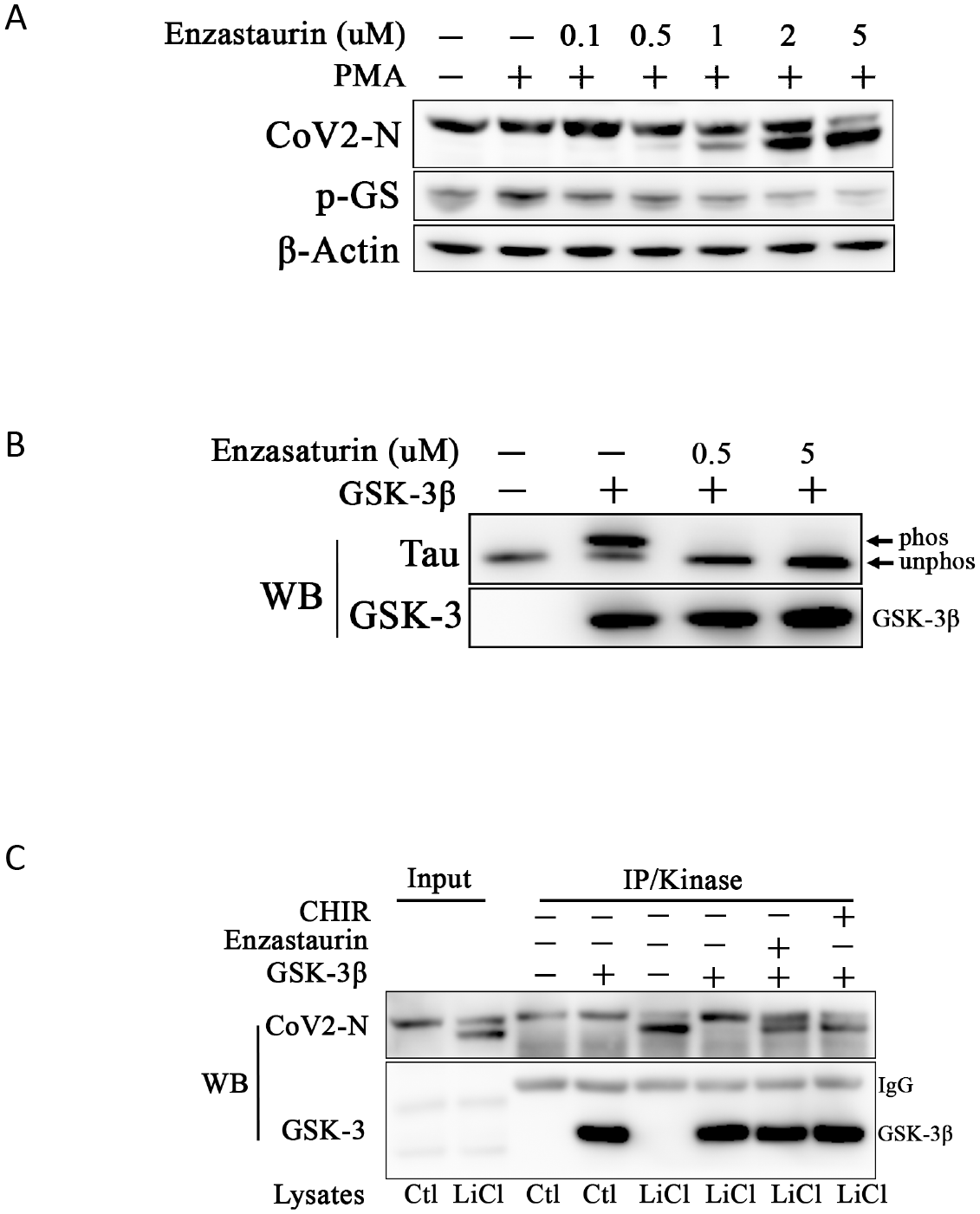
Enzastaurin inhibits N phosphorylation: **A.** N expressing 293T cells were treated with DMSO or increasing doses of Enzastaurin. Enzastaurin inhibited phosphorylation of N and GS in a dose dependent manner. Inhibition of PKC in these samples is described in supplemental figure S3. **B.** *In vitro* GSK-3 kinase assay using tau protein as substrate. Unphosphorylated tau migrates more rapidly (“unphos”) than tau phosphorylated by GSK-3 (“phos”). Enzastaurin inhibits GSK-3 activity directly at 0.5 µM. **C.** N protein was immunoprecipitated from HEK293T cells treated with 20 mM LCl as in Fig. 2D and added to an *in vitro* kinase reaction with recombinant GSK-3β. Phosphorylation of N protein was inhibited in the presence of Enzastaurin (10 µM) and CHIR99021 (2 µM).

### GSK-3 inhibitors block replication in SARS-CoV-2 infected cells

The GSK-3 inhibitors CHIR99021 and Enzastaurin were tested for antiviral efficacy and for their effects on cell viability at two institutions in two lung epithelial cell lines (Calu-3 and A549-Ace2). Calu-3 cells were treated with drugs at varying concentrations for 1 hour, inoculated with SARS-CoV-2, and cell number and the percent of infected cells were quantified at 48 hours post infection (HPI). CHIR99021 inhibited SARS-CoV-2 infection with an IC_50_ ∼5uM in Calu-3 cells (Fig. 4A), with marked reduction in frequency of infected cells at 10 µM (Fig. 4B). GSK-3 inhibition also reduced viral titers over 15-fold in the supernatant from SARS-CoV-2 infected Calu-3 cells (Fig. 4C). Furthermore, accumulation of phosphorylated N protein in infected Calu-3 cells, detected at 24-48 hours, was completely blocked by GSK-3 inhibition (Fig. 4D). CHIR99021 inhibition of SARS-CoV-2 in Calu-3 cells was similar in laboratories at the University of Pennsylvania and UCLA. CHIR98014, a GSK-3 inhibitor that is structurally similar to CHIR99021, was also reported to inhibit SARS-CoV-2 in A549-Ace2 cells at 5 µM (1), although we did not observe this effect in A549-Ace2 cells. Similarly, previous work has shown that Enzastaurin inhibits SARS-CoV-2-mediated cytopathic effect in Vero E6 cells at 250 nM(5) and reduces infection (based on qRT-PCR and viral titer) in A549-Ace2 cells at 5 µM (1); however, we did not observe an effect of Enzastaurin in A549-Ace2 or Calu-3 cells. The reasons for the cell-type variability in these assays is unclear but has been observed by others as well (1).

**Fig. 4.**
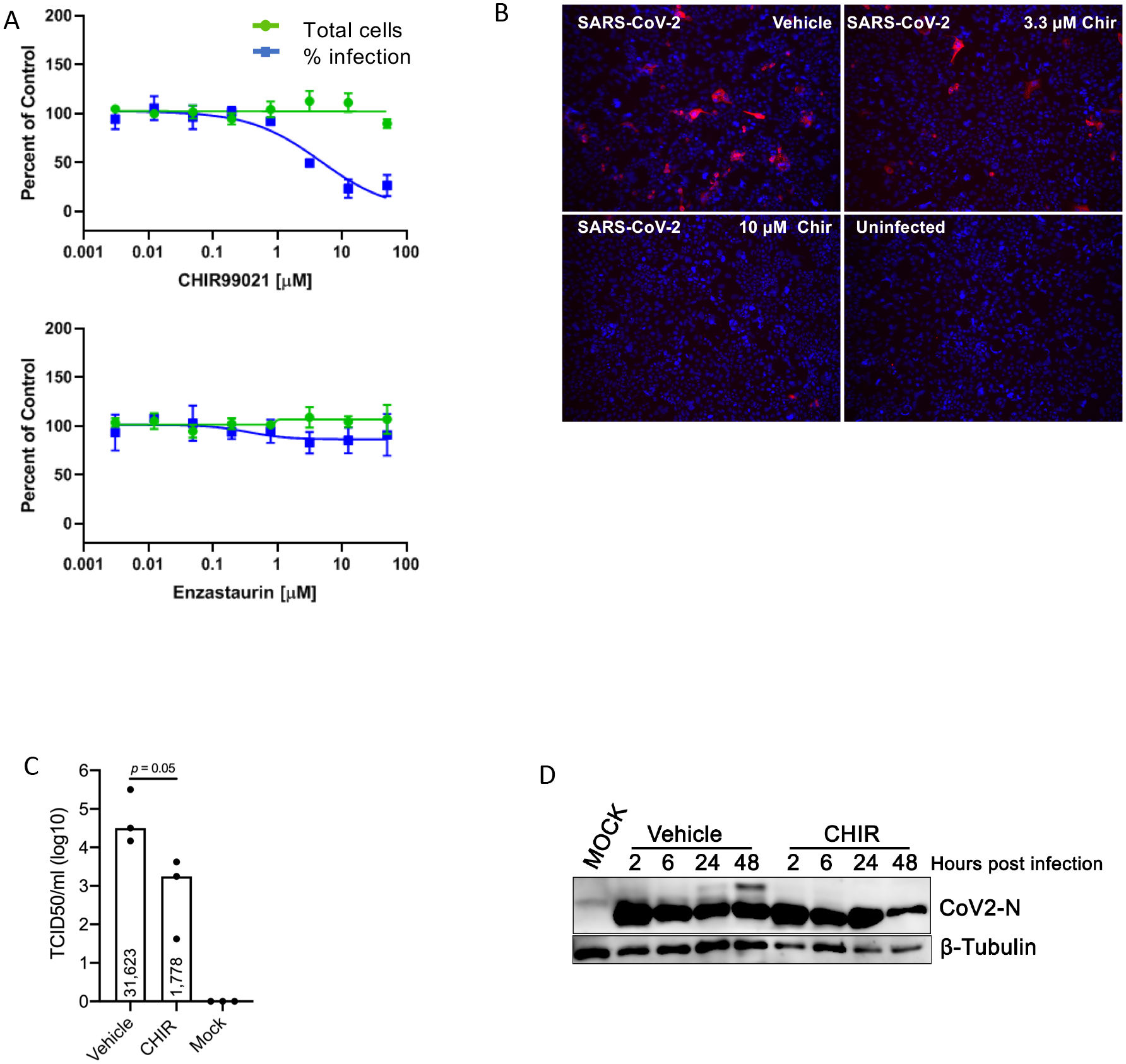
GSK-3 inhibitor blocks replication in SARS-CoV-2 infected cells: **A.** Dose-response analysis of Calu-3 cells treated with GSK-3 inhibitors CHIR99021 or Enzastaurin (UPenn). Cells were treated with drug at the indicated concentrations and then inoculated with SARS-CoV-2. Cells were fixed at 48hpi and total cell count (green) and percent viral infection (blue) detected by immunofluorescence for dsRNA were assessed. **B.** Calu-3 cells were treated with vehicle or the indicated concentrations of CHIR99021, innoculated with SARS-CoV-2, fixed at 48 hpi, and Spike protein was detected by immunofluorescence (UCLA). Enzastaurin had no effect on viral infection in Calu-3 cells. **C.** Calu-3 cells were treated with vehicle or CHIR99021 (10 µM), innoculated with SARS-CoV-2 at t = 0, and supernatants were sampled at 48 hpi for TCID50 quantification. Median titers are indicated within the boxes (note log_10_ scale). p = 0.05 (one tailed Mann-Whitney test). **D.** Calu-3 cells were treated with vehicle or CHIR99021, innoculated with SARS-CoV-2 at t = 0, and cell lysates were harvested for immunoblotting for N protein (upper panel) or tubulin (lower panel) at the indicated times after infection.

### Reduced risk of COVID-19 in lithium-treated patients

As lithium is a GSK-3 inhibitor that is widely used to treat bipolar disorder, we asked whether patients on lithium have a reduced risk of COVID-19 infection compared to the general population. We included patient data from three health systems in the United States (Fig. 5): 162,118 individuals from the University of Pennsylvania Health System (UPHS), 115,073 from Mount Sinai Medical Center (MSMC), and 102,420 from the University of Iowa Hospitals and Clinics (UIHC) who were tested for COVID-19 by RT-PCR (Table 1) as of February 2021. Among these participants, 13,641 (8.4%) patients had confirmed positive tests at UPHS, 10,597 (9.2%) at MSMC, and 16,170 (15.8%) at the UIHC. Across the three health systems, 7% of patients taking lithium developed COVID-19 compared with 15% among the general population. The average age of the patients who received lithium was between 42 – 48 years.

**Fig. 5.**
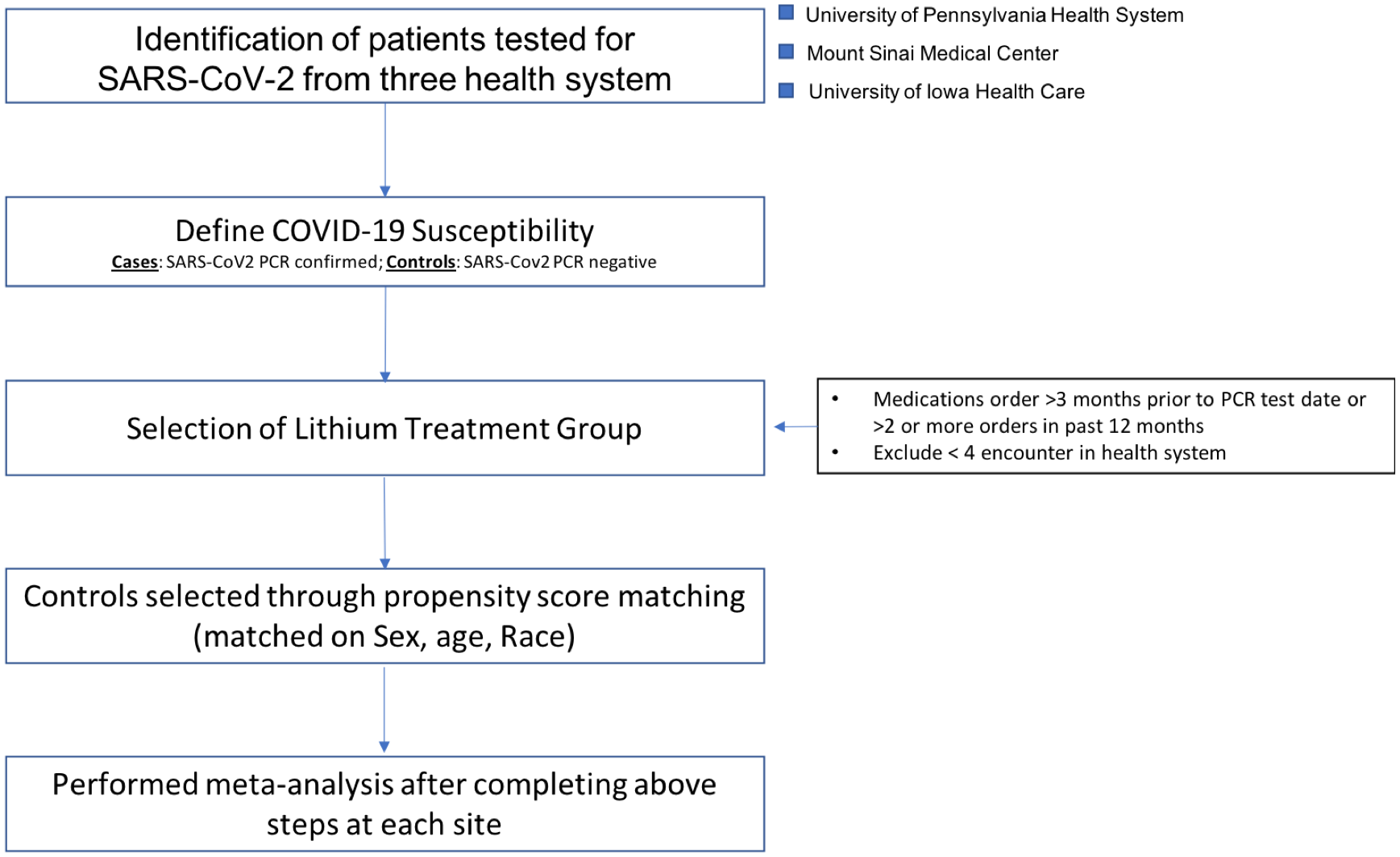
Overview of EHR analysis workflow. The flow diagram depicts the steps for sample selection, quality control and statistical approach to study assocciation between lithium and COVID-19 susceptibility using electronic health records.

**Table 1.**
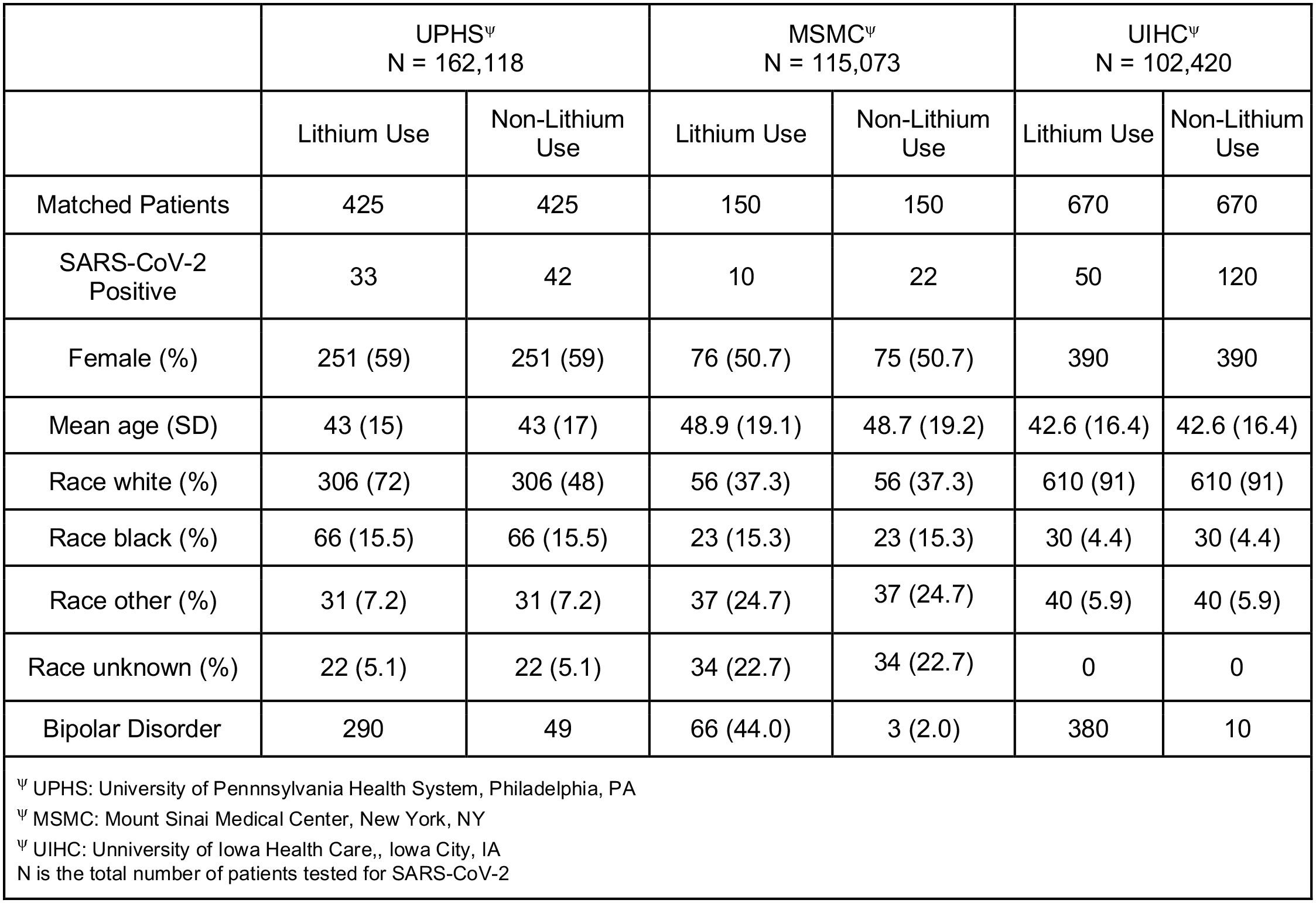
Patient characteristics of propensity score matched cohort from three health care systems.

Given the potential confounding bias for COVID-19 susceptibility with the patient’s baseline characteristics, propensity score matching was employed at each site. The matched cohort resulted in 33 patients from UPHS, 50 patients from UIHC, and 10 patients from MSMC who were on lithium treatment and tested positive for COVID-19 (Table 1). To increase the statistical power, we pooled the data from three sites by conducting a meta-analysis using a random effects model (Table 2). We found that patients on lithium had reduced risk of COVID-19 infection compared with nonusers (p-value= 0.005, OR=0.51 [0.35 - 0.74]). The test for heterogeneity had low variance between the observations from three sites (Q statistic P = 0.12, I^2^ = 51.6%). Additionally, a subgroup analysis among patients with bipolar disorder taking lithium compared to nonusers of lithium was conducted. The association was not statistically significant (p > 0.01). There were considerably fewer BPD patients tested for COVID-19 in our study population which may have led to low statistical power to detect association.

**Table 2.** Association between lithium use and COVID-19 susceptibility.

| Study Cohort | Lithium<br>(n/N) | Control group<br>(n/N) | Relative Risk<br>95% CI |
| --- | --- | --- | --- |
| University of Pennsylvania Health System | 33/435 | 46/435 | 0.71 [0.46 - 1.09] |
| Mount Sinai Medical Center | 10/150 | 22/150 | 0.45 [0.22 - 0.92] |
| University of Iowa Health Care | 50/670 | 120/670 | 0.42 [0.31 - 0.57] |
| Meta-analysis* |  |  | **0.51** [0.35 - 0.74] |
| * Meta-analysis statistics<br>Test for random effect $z = -3.47$, $p = 0.005$<br>Test for heterogeneity $df = 2$, $p = 0.12$, $I^2 = 51.6\%$<br>N = Total number of patients in the cohort<br>n = Number in group with outcome |  |  |  |

## Discussion

Medications that target common features of the coronavirus family could reduce the severity and transmission of COVID-19 as well as other pathogenic coronaviruses. Our analysis of retrospective EHR data on SARS-CoV-2 PCR testing from three major health systems across the US showed a ∼50% reduced risk of COVID-19 in patients taking lithium. We show that the SARS-CoV-2 N protein is phosphorylated by GSK-3 and that lithium and other GSK-3 inhibitors block N phosphorylation, as shown previously for JHMV and SARS-CoV(9). We also show for the first time that GSK-3 is unequivocally essential for N phosphorylation using *GSK3A/B* double KO. GSK-3 inhibition blocks phosphorylation of virally encoded N protein in SARS-CoV-2 infected human cells, reduces intracellular viral RNA accumulation, and reduces viral titers in cell culture supernatants. GSK-3 inhibition may therefore allow safe and effective therapy for COVID-19. As we find GSK-3 consensus sites in the N proteins of diverse coronaviruses, GSK-3 inhibitors may also be effective antiviral therapy in other coronavirus infections, including those that may arise in the future.

Lithium has a narrow therapeutic window, however, and the concentration needed to inhibit N phosphorylation and to impair infectivity of SARS-CoV-2 and other coronaviruses in cell culture(7, 9, 19–21) is above the therapeutic range in humans (peak level ∼2 mM, trough levels ∼1 mM(17)) and the level that induces behavioral changes in model organisms. For example, 1 mM lithium is sufficient to inhibit GSK-3 in humans(34), alter GSK-3 dependent behaviors in mice (33, 45), and activate Wnt signaling in the mouse intestine(46, 47), whereas the IC_50_ for inhibition of N phosphorylation and inhibition of SARS-CoV-2 replication, as well as inhibition of phosphorylation of established GSK-3 substrates, in cultured cells is ∼10 mM. It is not clear why higher concentrations were needed to inhibit GSK-3 in cell culture compared to in vivo studies, but, given the narrow therapeutic window for lithium in BD, alternative GSK-3 inhibitors may be better tolerated in clinical settings involving coronavirus infection.

While the association of lithium therapy and reduced risk of COVID-19 across three health systems is both signficant and intriguing, observational studies have many limitations. A variety of factors with potential biases cannot be measured even after comparing the cases and controls in a manner that accounts for known confounding factors using a rigorous matching algorithm. For instance, details on medicine usage were derived from records of prescription orders, but information on compliance before SARS-CoV-2 PCR testing is not available. In addition, the collection of a non-random sample population can create a collider bias and lead to distorted associations. For example, the COVID-19 test was restricted particularly in the early pandemic to symptomatic patients so that many asymptomatic patients in the EHR were not tested. These findings should therefore be interpreted carefully and deeper investigation is required in a cohort with a larger sample size.

Prior work has shown that lithium and the GSK-3 inhibitor Kenpaullone inhibit N phosphorylation and reduce viral titers in SARS-CoV and JHMV infected Vero6 cells(8, 9) and *GSK3* knockdown also impairs replication of IBV in Vero cells(15). We also show that multiple small molecule GSK-3 inhibitors, including CHIR99021, BIM-I, AR-A014418, Enzastaurin, and Sotrastaurin block SARS-CoV-2 phosphorylation. These pharmacological studies are compelling evidence that GSK-3 is a critical host kinase for N protein, but these drugs may have off-target effects. Thus, the complete abrogation of N phosphorylation by the double KO of *GSK3A* and *GSK3B* demonstrates unequivocally that GSK-3 is essential for phosphorylation of N at these sites.

The highly selective GSK-3 inhibitor CHIR99021 impaired N phosphorylation and reduced SARS-CoV-2 infection in the human lung epithelium-derived cell line Calu-3, an observation that was reproducible in two independent laboratories, and the related compound CHIR98014 was previously reported to inhibit infection in the human lung cancer derived cell line A549-Ace2(1). Furthermore, the clinically well tolerated drug Enzastaurin was reported to inhibit SARS-CoV-2 infection in A549-Ace2 cells(1) and viral-mediated cytopathic effect in Vero E6 cells(5). However, the effects of these inhibitors has been variable in different cell lines and in different laboratories. For example, Bouhaddou et al did not observe inhibition with Enzastaurin in Vero6 cells and we did not observe inhibition with Enzastaurin in A549-Ace2 cells or Calu-3 cells. The reasons for this cell type specific effect and variability between laboratories is unclear, but may include differences in the expression and/or activity of targeted signaling pathways in different cell lines that may arise as an adaptation to cell culture conditions and passage number, as well as differences in infection time or assays used to assess infection. Nevertheless, it remains clear that GSK-3 is essential for N phosphorylation, as *GSK3* knockout abrogates N phosphorylation, and given the essential functions of phosphorylated N in viral transcription, replication, and packaging(8), developing GSK-3 inhbitors that safely and effectively inhibit N phosphorylation is a promising potential approach to controlling SARS-CoV-2 and other coronavirus infections that may arise in the future.

We propose that inhibition of N phosphorylation underlies the antiviral activity of lithium and other GSK-3 inhibitors; however GSK-3 also regulates inflammatory responses (48), and lithium has been reported to have antiviral activity against other viruses, notably human herpes viruses (22). Thus modulation of the inflammatory response by lithium may also contribute to the reduced risk of COVID-19 in patients taking lithium.

The search for antiviral drugs that target coronaviruses is progressing rapidly. Multiple bioinformatic analyses and high throughput screens have identified an array of promising candidates with efficacy in cultured cells (1–4, 49). Our approach is based on a clear mechanism, utilizes clinically-tested and well-tolerated drugs that could be rapidly repurposed for COVID-19, and is supported by clinical data showing an association between treatment with a GSK-3 inhibitor (litihum carbonate) and reduced risk of COVID-19. Inhibition of a host protein required for coronavirus propagation has a potential advantage over inhibiting viral proteins as the probability of developing drug resistant mutations should be lower for the host protein. Given the major coronavirus epidemics over the past 20 years, including SARS, COVID-19, and MERS, new coronavirus outbreaks in the future are possible. Multiple approaches will be needed to address current and potential future outbreaks, including the development of medications that target common features of coronavirus. Interfering with the conserved dependence of the nucleocapsid protein on the host protein GSK-3 may be a viable approach to treat COVID-19 and potential future coronavirus outbreaks.

## Methods

### Plasmids, antibodies

The SARS-CoV-2 nucleocapsid gene was PCR amplified from cDNA derived from the isolate SARS-CoV-2/human/USA/CA-CZB017/2020 (Genbank MT385497.1), nucleotides 28254-29513, and cloned into pCS2MT in frame with five N terminal myc epitope tags (https://www.addgene.org/153201/). Priming site mutations were generated by site directed mutagenesis to modify serine-188 and serine-206 to alanine based on prior observations with SARS-CoV-1(9). Antibodies to the SARS-CoV-2 N protein were purchased from Invitrogen (#PA1-41386). Antibodies from Cell Signaling included phospho-Glycogen Synthase (#3891), phospho-S6 (#4858), β-catenin (#9562), phospho-β-catenin (#9561), GAPDH (#2118), PKCα (#2056), PKCδ (#2058), PKCε (#2683), Myc-tag (#2276), and phospho (Ser) substrate (#2261). Other antibodies included antibodies to GSK-3 (Calbiochem #368662), Tau (T14/46 antibodies provided by Virginia Lee, University of Pennsylvania), and β-actin (Sigma #A5441). Monoclonal Anti-SARS-CoV S Protein (similar to 240C) was obtained through BEI Resources, NIAID, NIH (NR-616).

### Cell culture, transfections, and CRISPR/Cas9 knockout

HEK293T cells (ATCC #CRL-1573) were cultured in Dulbecco’s Modified Eagle Medium (DMEM, GIBCO #11965) supplemented with 10% Fetal Bovine Serum (Hyclone #SH30071.03) and 1% penicillin/streptomycin (GIBCO #15140), and were maintained at 37°C and 5% CO2. Cells were transfected 24 hours after plating using Lipofectamine 3000 (Invitrogen #L3000001) for plasmids or Lipofectamine RNAiMax (Invitrogen #13778075) for siRNAs according to manufacturer’s instructions. For silencing PKC isozymes, siRNA oligonucleotides from Dharmacon (Lafayette, CO) were: J-003523-17-0002 (PKCα), J-003524-08-0002 (PKCδ), and J-004653-08-0002 (PKCε). Transfections were done in Opti-MEM Reduced Serum Medium (GIBCO #31985). For CRISPR/Cas9 knockout, guide RNA (gRNA) sequence targeting GSK-3A (GCCTAGAGTGGCTACGACTG) or GSK-3B (AGATGAGGTCTATCTTAATC) was cloned into lentiCRISPRv2 vector (Addgene #99154, Sanjana et al. 2014). Lentivirus was packaged as previously described (Hou, et al. 2015). For transduction, 2X10^5^ 293T cells were seeded in 6-well plate with 10ug/ml polybrene and transduced with the lentivirus at a multiplicity of infection (MOI) of 20. After 24 hours, medium was changed. Two days after transduction, about 1,000 tranduced 293T cells were mixed with 1ml of methylcellulose (MethoCult H4034 Optimum, Stem Cell Technologies) into a 6-well plate and cultured at 37°C and 5% CO2 for two weeks. Single-clone colonies were then picked and cultured in 96-well plate. The cells were passaged every 2-3 days, and half of the cells was collected for genomic DNA extraction. Then GSK-3 target region was PCR amplified and sequenced. GSK-3A PCR primers: forward, GTCCCAGCATCCACCTTTCCTCA; reverse, ACCTGAGTTTGTTTCCCTGCTTT. GSK-3B PCR primers: forward, ATAGGATATGAGGACATTGAT; reverse: GCAGAAATAAAATCTATAAATGTCTGTG. Immunoblot analysis was performed to confirm knockout of single clones.

### Immunoblotting, immunoprecipitations, and *in vitro* protein kinase assay

Cells were lysed in buffer containing 20mM Tris pH 7.5, 140mM NaCl, 1mM EDTA, 10% glycerol, 1% Triton X-100, 1mM DTT, 50mM NaF, and protease inhibitor cocktail (Sigma P8340), phosphatase inhibitor cocktail #2 (Sigma P5726) and #3 (Sigma P0044) used 1:100 each. Supernatants were collected after centrifugation at 14,000 rpm for 15 min at 4°C, adjusted to 1x laemmli sample buffer and subjected to SDS-PAGE and immunoblotted as described previously(9). To resolve phosphorylated forms of Nucleocapsid protein, cells were lysed in the above lysis buffer and twenty micrograms of protein lysate was electrophoresed on 10% NuPAGE Bis-Tris polyacrylamide gels (Invitrogen, #NP0301) for 17 hours at 70 V and then subjected to immunoblotting as above(9). For immunoprecipitation/protein kinase assays, anti-Myc-tag antibody was incubated with Surebeads Protein G megnetic beads (Bio-Rad, #1614023) for 10 min, lysates were then added for an additional 1 hr at room temperature. Antibody-bound beads were washed with PBS-T (PBS + 0.1% Tween 20) 3 times and resuspended in 1X protein kinase buffer (100mM Tris pH 7.5, 5mM DTT, 10mM MgCl_2_, 60uM ATP, ± human recombinant GSK-3β) and incubated at 30°C for 30 min. Reactions were stopped by adding standard 2X Laemmli Sample Buffer and incubating at 70°C for 10 min and then subjected to SDS-PAGE and immunoblotting as above. ^32^P labeling was performed similarly except that *γ*-^32^P-ATP was added (∼10 µCi) to each reaction and samples were resolved by SDS-PAGE and autoradiography.

### Infection with SARS-CoV-2

UPenn: SARS-CoV-2 (Isolate USA-WA1/2020) was obtained from BEI Resources. Stocks were prepared and titered (1×10^7^ pfu/mL and 1.5×10^6^ TCID50/ml) as described previously(2). Cells were plated in 384-well plates (20 µl/well) at 7500 cells per well. The next day, 50 nL of drugs or vehicle control (DMSO) were added and one hour later cells were inoculated with SARS-CoV-2 (MOI=0.5) and incubated for 48 hours. Cells were then fixed in 4% formaldehyde/PBS, immunostained for dsRNA (anti-dsRNA J2), and imaged as described previously(2). The total number of cells and the number of infected cells were measured using MetaXpress 5.3.3 cell scoring module and the percentage of infected cells was calculated. All work with infectious virus was performed in a Biosafety Level 3 laboratory and approved by the Institutional Biosafety Committee and Environmental Health and Safety.

UCLA: Calu3 cells were seeded at 3×10^4^ cells per well in 0.2 ml volumes in 96-well plates. CHIR99021 or Enzastaurin (each at 10 µM and 3.3 µM) was added to cells followed after 1 hour by inoculation with SARS-CoV-2 (isolate USA-WA1/2020) at a multiplicity of infection (MOI) of 0.1. At 48 hours post infection (hpi), the cells were fixed with 4% paraformaldehyde and viral infection was examined by immunofluorescent analysis (IFA) using SARS-CoV Spike (S) antibody [BEI Resources: NR-616 Monoclonal Anti-24 SARS-CoV S Protein (Similar to 240C)]. For immunoblotting of infected cells, cells were lysed in RIPA buffer at different times as indicated in Fig. 4D and subjected to SDS-PAGE (NuPAGE) and immunoblotting as above. Viral titer was quantified in cell culture supernatants using Vero E6 cells and TCID50 was determined as described(5).

### Study design and cohort selection for electronic health records (EHR) analysis

We extracted retrospective EHR data on patients with a documented SARS-CoV-2 infection from three major health care systems across the United States. The cohort included patients from the University of Pennsylvania Health System (UPHS), Mount Sinai Medical Center (MSMC), and University of Iowa Health Care (UIHC). Our main hypothesis was to test for the association between lithium use and COVID-19 susceptibility. We employed several EHR phenotyping algorithms to identify patients who were tested for COVID-19 by RT-PCR, patients with prescriptions for lithium, and patients with bipolar disorder. To account for the possibility that an individual had been tested for COVID-19 within the health system, but otherwise managed their care at a different health system, such that they had no medication or diagnosis information, we restricted the analysis to individuals with at least four encounters within the health system. The analysis workflow is provided in Fig. 5. The full study protocol for UPHS EHR analysis was approved by the University of Pennsylvania Institutional Review Board (IRB) under the protocol for the study titled “Clinical, social, and genetic risk stratification for COVID-19 outcomes” (Protocol #844360) and for MSMC under the protocol titled “MSCIC Predictive Modeling & Consultation Tool” (IRB Protocol #20-00338). Additionally, under Penn Medicine’s HIPPA notice of privacy practices, patients consent to use of EHR data for research study authorized by University of Pennsylvania Institutional Review Boards. All the data used in the analysis was deidentified. Hence, explicit consent from the patients was not required. Link to HIPAA notice: https://www.pennmedicine.org/for-patients-and-visitors/patient-information/hipaa-and-privacy/hipaa-notice-of-privacy-practices.

### Outcome

The primary outcome of our case-control EHR study was COVID-19 susceptibility where cases are defined by positive test results from RT-PCR of nasal samples and controls with negative test results. PCR test results were recorded through February, 2021. The analysis was also restricted to participants between the ages of 18 and 89.

### Exposure

Lithium use was defined using the prescription orders available within the EHR. The medication name and dose generally differ across health systems and it poses a challenge to develop standard selection criteria. To minimize these differences, we used RxNorm – a resource of standardized nomenclature for drug names from the National Library of Medicine(50). RxNorm maps branded and generic names, ingredients, drug components, and other drug-related vocabularies to standard names. The current EHR systems (EPIC) also support RxNorm and there is an existing mapping between drug names and RxNorm concept unique identifiers (RxCUI). We queried RxNorm to extract all the RxCUI linked with lithium carbonate and then extracted prescription orders mapped to RxCUI in the EHR system. The list of RxNorm CUIs can be found in Supplementary Table 2. A patient was considered on lithium treatment if they had an order placed within 90 days prior to their first positive COVID-19 test (COVID-19 cases) or 90 days before their first negative COVID-19 test (COVID-19 controls). Generally, lithium is prescribed for a longer period of time (> 3 months), so to capture long-term use of lithium we included patients with two or more lithium orders placed within 12 months before their COVID-19 test, using the aforementioned methods for COVID-19 cases and controls.

### Statistical Analysis

To minimize potential confounding biases among the population tested for COVID-19, we applied a propensity score matching (PSM) method(51). For each patient on lithium with a record of COVID-19 testing, we first calculated the propensity score using a multivariate logistic regression model adjusting for age, sex, and race. Then, we applied nearest-neighbor matching (*MatchIt* R) on the propensity scores to select one matched patient for each patient on lithium. We conducted a meta-analysis on the association between lithium use and COVID-19 outcome occurrences using the *meta* R package. We pooled the effect estimates using fixed effect and random effects models and the Mantel-Haenszel method was used to calculate the fixed effect estimate. Since we have effect estimates from only three sites, a sensitivity analysis of subgroups was not feasible. We assessed the heterogeneity of the meta-analysis through I^2^ and Chi^2^ statistics. TCID50 data (vehicle versus CHIR) were analyzed for statistical significance using non-parametric t test (one-tailed Mann-Whitney test) with GraphPad Prism software, version 8.2.1 (GraphPad Software, US).

## Data Availability

Data are available upon request from the authors. Electronic Health Record data are available upon request from qualified investigators subject to a data use agreement and IRB approval.

## Acknowledgements

The following reagents were obtained through BEI Resources, NIAID, NIH: Monoclonal Anti-SARS-CoV S Protein (Similar to 240C), NR-616 and SARS-Related Coronavirus 2, Isolate USA-WA1/2020, NR-52281, which was depostied by the Centers for Disease Control and Prevention. Funding: PSK is supported by grants from the National Institutes of Health (R21AI161678, 1R01HL141759, 1R01GM115517). VA is supported by the UCLA DGSOM and Broad Stem Cell Research Center institutional award (OCRC #20-15), National Institute of Health (1R01EY032149) and the California Institute for Regenerative Medicine TRAN Award (TRAN1COVID19-11975). We thank the Dean’s Innovation Fund, Linda and Laddy Montague, BWF, NIAID (5R01AI140539, 1R01AI1502461, R01AI152362), NCATS, the Fast Grants Award from Mercatus, and the Bill and Melinda Gates Foundation for funding to SC. SC is an Investigator in the Pathogenesis of Infectious Diseases from the Burroghs Wellcome Fund.

## Author contributions (CRediT statement)

Conceptualization: PSK, XL, AV

Methodology: XL, AV, HR, DS, SC, RD, VA, MGK

Formal Analysis: AV, JJM, AK

Investigation: XL, AV, HR, GG, RM, AL, WC, JJM, AC, AK, MGK, DJR, MDR, WHB, RD, VA, DS, SC, PSK

Resources: WC, AC Visualization: XL, AV, SC, RD, VA

Funding acquisition: PSK, VA, SC, MDR

Supervision: WC, AC, DJR, MDR, WHB, RD, VA, SC, PSK

Writing – original draft: PSK, XL, AV

Writing – review & editing: XL, AV, VA, MDR, PSK

## Competing Interests Statement

The authors declare no competing interests. MDR is on the Scientific Advisory Board for Goldfinch Bio and Cipherome.

## Supplementary Figures and Table

### Supplementary Figures

Fig. S1. N phosphorylation is blocked by GSK-3 inhibitors.

Fig. S2. GSK-3 phosphorylation of N requires priming site but is independent of mTORC1 or SRPK.

Fig. S3. Enzastaurin and Sotrastaurin block N phosphorylation through inhibition of GSK-3.

**Supplementary Figure S1.**
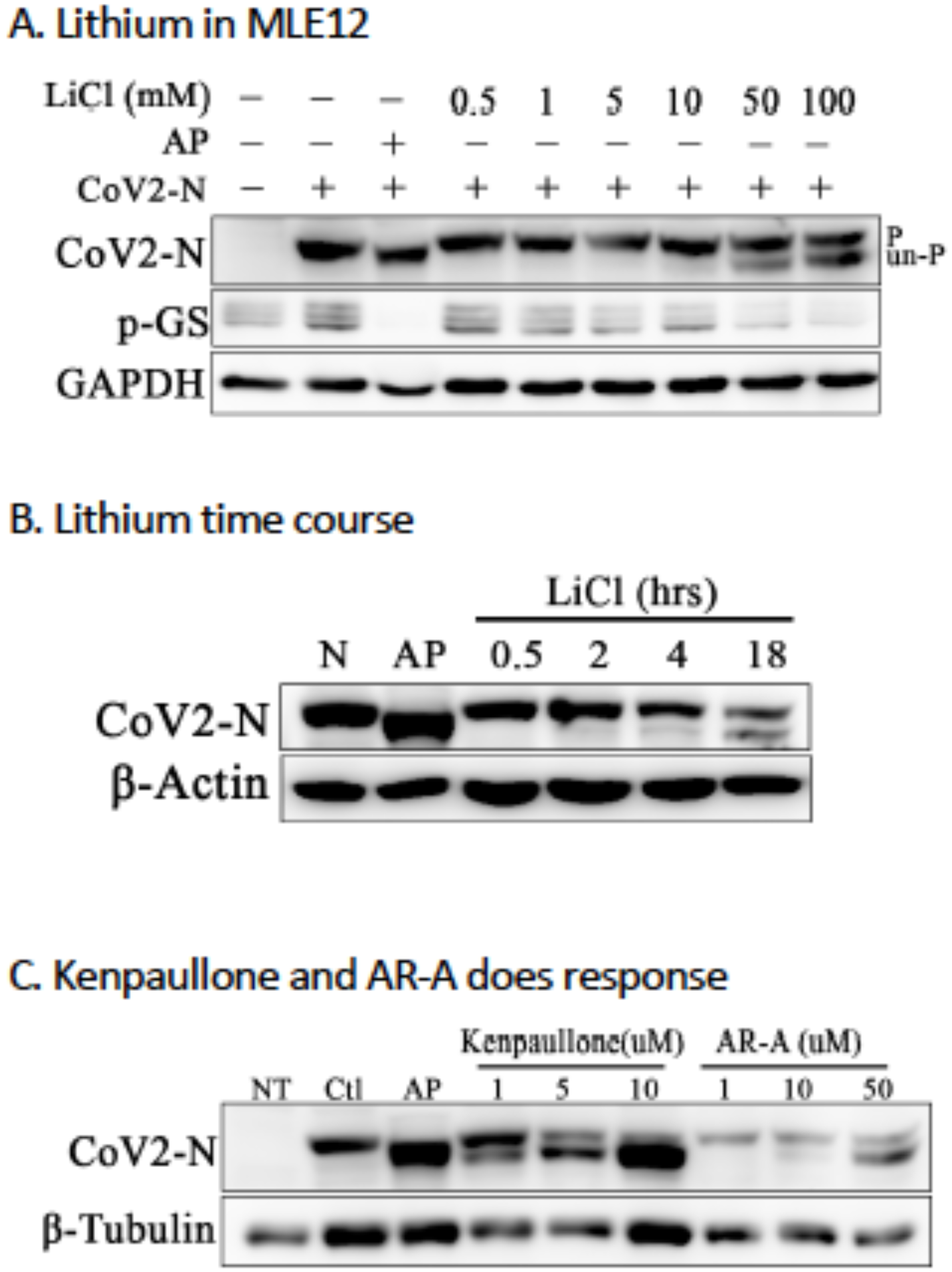
**A.** SARS-CoV-2 N was expressed in MLE12 cells, treated with LiCl for 18h, and then subjected to SDS-PAGE and immunoblotting for N protein (upper panel) and phosphorylated β-catenin (lower panel). Alkaline phosphatase (AP) treatment of cell lysates increases electrophoretic mobility. “P” indicates slower migrating, phosphorylated N and “un-P” indicates dephosphorylated species. **B.** N was expressed in 293T cells, treated with 10 mM LiCl, and cell lysates were harvested at different time points for Western blot analysis. CoV-2 N protein (upper panel); β-actin (lower panel) is loading control. **C.** Alternative GSK-3 inhibitors Kenpaullone and AR-A014418 inhibit N protein phosphorylation. SARS-CoV2 N expressing 293T cells treated for 18h with Kenpaullone or AR-A014418 at the indicated concentrations were immunoblotted as in panel B.

**Supplementary Figure S2.**
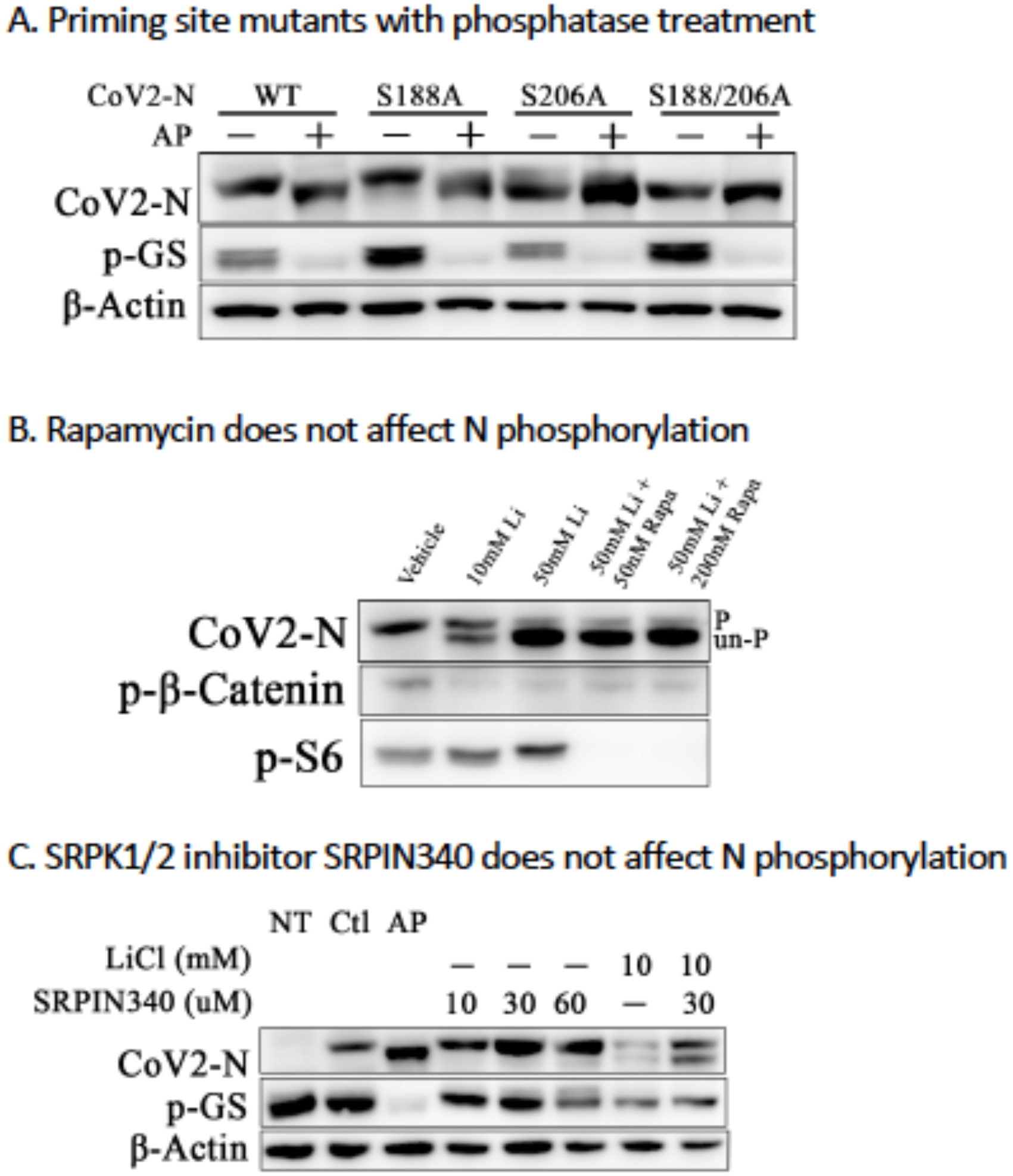
**A.** Serine-188 and serine-206 of SARS-CoV-2 N protein were mutated to alanine and single and double mutant N proteins were expressed in 293T cells. Alkaline phosphatase (AP) treatment of cell lysates increases electrophoretic mobility. **B.** mTOR inhibitor Rapamycin had no effect on N phosphorylation. SARS-CoV2 N expressing 293T cells were treated with Rapamycin (50 and 200 nM) in the presence of LiCl for 18h and then subjected to SDS-PAGE and immunoblotting for N protein (upper panel) and phosphorylation of β-catenin (middle panel) and S6 (lower panel). Rapamycin inhibited S6 phosphorylation but had no effect on N phosphorylation. “P” indicates phosphorylated N; “Un-P” indicates dephosphorylated species. **C**. N expressing 293T cells were treated with the SRPK inhibitor SRPIN340 or LiCl separately or in combination and cell lysates were then harvested for Western blot analysis as in panels A and B.

**Supplementary Figure S3.**
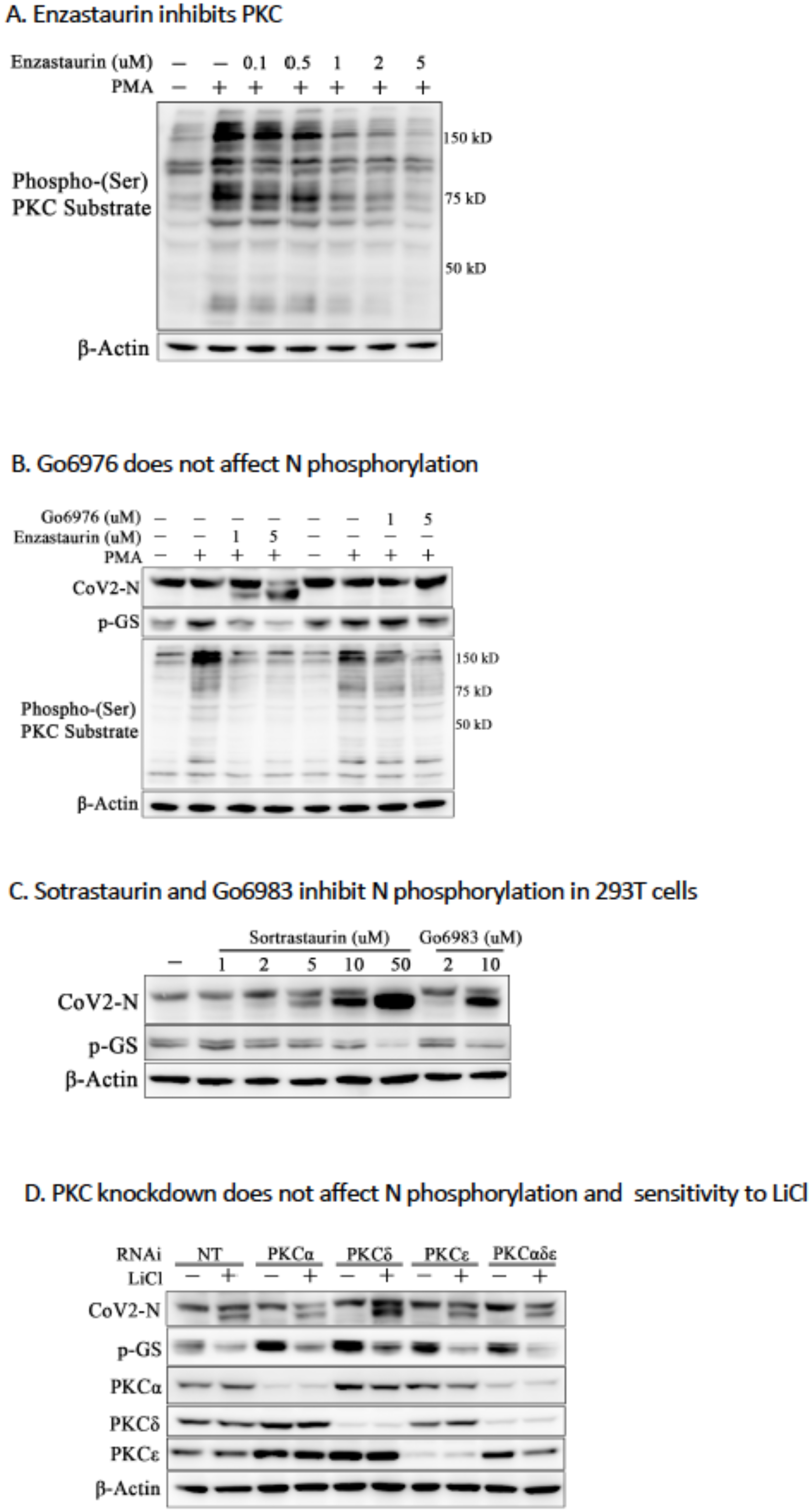
**A.** N expressing 293T cells were treated with DMSO or increasing doses of Enzastaurin and then stimulated with 200 nM phorbol 12-myristate 13-acetate (PMA) for 30 mins prior to lysis. PKC activity was assessed with an antibody that detects phosphorylated PKC substrates. β-actin is shown as a loading control. **B.** N expressing 293T cells were treated with DMSO or increasing doses of the PKC inhibitors Enzastaurin and Gö6976 and stimulated with PMA as in panel A. Cell lysates were immunoblotted for N protein (upper panel), phospho-GS (middle panel), or phosphorylated PKC substrates (lower panel). Gö6976 does not affect phosphorylation of N or GS. **C.** The structurally related PKC inhibitors Sotrastaurin and Gö6983 inhibit N phosphorylation. N expressing cells were treated with Sotrastaurin or Gö6983 at the indicated concentrations and then lysates were immunoblotted for N and p-GS as above. **D.** Global PKC knockdown does not affect phosphorylation of N protein or GS. N expressing 293T cells were treated with vehicle or LiCl, then transfected with siRNAs directed against *PKCα, PKCδ*, or *PKCε* individually or in combination, as described previously. Cell lysates were immunoblotted for CoV-2 N protein, pGS, or PKC isoforms as indicated in figure. β-actin is shown as a loading control.

**Supplementary Table 1.**
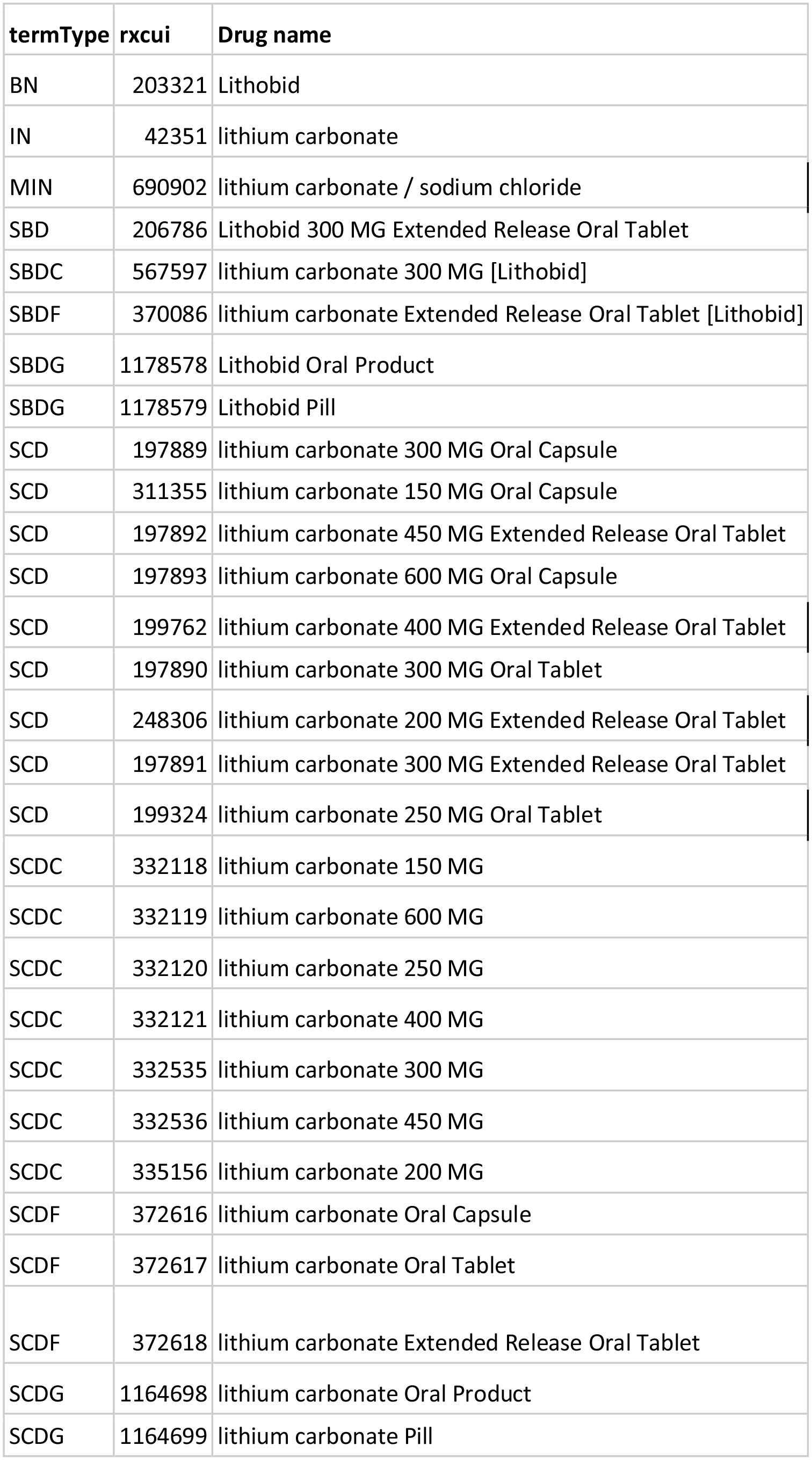
RxNorm concept unique identifiers (CUI) for lithium carbonate used in the EHR analysis.

## Notes

### Funding Statement

PSK is supported by grants from the National Institutes of Health (1R01HL141759 and 1R01GM115517). VA is supported by the UCLA DGSOM and Broad Stem Cell Research Center institutional award (OCRC #20-15), National Institute of Health (1R01EY032149), and the California Institute for Regenerative Medicine TRAN Award (TRAN1COVID19-11975). We thank the Dean’s Innovation Fund, Linda and Laddy Montague, BWF, NIAID (5R01AI140539, 1R01AI1502461, R01AI152362), NCATS, the Fast Grants Award from Mercatus, and the Bill and Melinda Gates Foundation for funding to SC. SC is an Investigator in the Pathogenesis of Infectious Diseases from the Burroghs Wellcome Fund.

### Author Declarations

EHR analysis was performed with Institutional Review Board (IRB) approval from the University of Pennsylvania (IRB #844360) and the Mt. Sinai Medical Center (IRB #20-00338).

### Summary of Updates

Expanded data on effect of GSK-3 inhibition on SARS-CoV-2 infected cells. Improved resolution of SDS-PAGE. P-32 labeling of N by GSK-3.

